# Detection of SARS-CoV-2 in Exhaled Breath from COVID-19 Patients Ready for Hospital Discharge

**DOI:** 10.1101/2020.05.31.20115196

**Authors:** Lian Zhou, Maosheng Yao, Xiang Zhang, Bicheng Hu, Xinyue Li, Haoxuan Chen, Lu Zhang, Yun Liu, Meng Du, Bochao Sun, Yunyu Jiang, Kai Zhou, Jie Hong, Na Yu, Zhen Ding, Yan Xu, Min Hu, Lidia Morawska, Sergey A. Grinshpun, Pratim Biswas, Richard C. Flagan, Baoli Zhu, Wenqing Liu, Yuanhang Zhang

## Abstract

The COVID-19 pandemic has brought an unprecedented crisis to the global health sector^1^. When recovering COVID-19 patients are discharged in accordance with throat or nasal swab protocols using reverse transcription polymerase chain reaction (RT-PCR), the potential risk of re-introducing the infection source to humans and the environment must be resolved^2,3,4^. Here we show that 20% of COVID-19 patients, who were ready for a hospital discharge based on current guidelines, had SARS-CoV-2 in their exhaled breath (∼10^5^ RNA copies/m^3^). They were estimated to emit about 1400 RNA copies into the air per minute. Although fewer surface swabs (1.3%, N=318) tested positive, medical equipment frequently contacted by healthcare workers and the work shift floor were contaminated by SARS-CoV-2 in four hospitals in Wuhan. All air samples (N=44) appeared negative likely due to the dilution or inactivation through natural ventilation (1.6–3.3 m/s) and applied disinfection. Despite the low risk of cross environmental contamination in the studied hospitals, there is a critical need for strengthening the hospital discharge standards in preventing re-emergence of COVID-19 spread.

## Main Text

The world has almost been brought to a standstill by the COVID-19 pandemic, while hospitals around the world are overwhelmed with unprecedented challenges^1^. Inside COVID-19 patient care centers, the air, frequently touched object surfaces, and floors have been found to be contaminated by SARS-CoV-2^5,6,7^. Infections of medical staff by SARS-CoV-2 were reported in hospitals across the globe; some of them have unfortunately lost their lives^8,9^. Stressfully, the medical community is also coping with the dilemma of discharging patients to re-allocate precious resources; while accepting the risks of reintroducing the source of infection to the public. For convenient diagnosis of the disease, the hospitals use throat or nasal swabs, sometimes supplemented with computed tomography (CT) scans for enhanced screening^10,11^. After hospital discharge, some COVID-19 patients, however, have tested positive again for SARS-CoV-2 using the throat swab protocol^2,3,4^. Despite major progress in understanding the COVID-19, infection risks related to recovering patients and to contaminated surfaces and air are not well documented. In this work, we aimed to examine the possible presence of SARS-CoV-2 in exhaled breath specimens from recovering COVID-19 patients who have repeatedly tested negative using throat swabs; and to study environmental contamination by SARS-CoV-2 within four hospitals in Wuhan, China.

Here we show that the exhaled breath condensate (EBC) samples from recovering COVID-19 patients in Wuhan hospitals (Table S1) tested positive for SAR-CoV-2 RNA (Fig. 1). These patients were over 70 years old (Fig. 1A, Table S2, S3). The EBC samples were collected at least 14 days after they had developed clinical symptoms (Fig. 1B); their throat swabs repeatedly tested negative at the time of the EBC collection and analysis. Their IgG tests were yet positive on 6 and 9, March about 4–7 days before their EBC samples were collected (Fig. 1C). Surprisingly, for patient B-L1(A), all tests with the throat swabs since 21 February had been negative, but the patient’s EBC sample, which was collected on 13 March (about 36 days after the patient developed symptoms), tested positive for SARS-CoV-2. For patient B-Z1(B), throat swabs tested positive on 21 February, 2 March, and then negative on 11 March; of two tests performed on March 12, one tested positive, and the other tested negative. On the following days (13,14, and 15 March) the patient’s throat swabs, however, repeatedly tested negative (Fig. 1D). Similar to patient B-L1 (A), patient B-Z1’s EBC sample, which was collected on 13 March (at least 43 days after the patient developed clinical symptoms), tested positive. ORF1a/b genes for SARS-CoV-2 were not detected in either patient (Fig. 1C). The throat swabs from three non-COVID-19 patients all tested negative for SARS-CoV-2. For one non-COVID-19 patient (C-Y1), two EBC samples were collected, all of which tested negative by RT-PCR. The EBC samples of one COVID-19 subject (A-X1) and two non-COVID-19 subjects (A-J1 and A-U1) also tested negative using the chip technology as described in the Methods. Overall, the EBC sample positive rate was 20% among the 10 recovering COVID-19 patients.

**Fig. 1.**
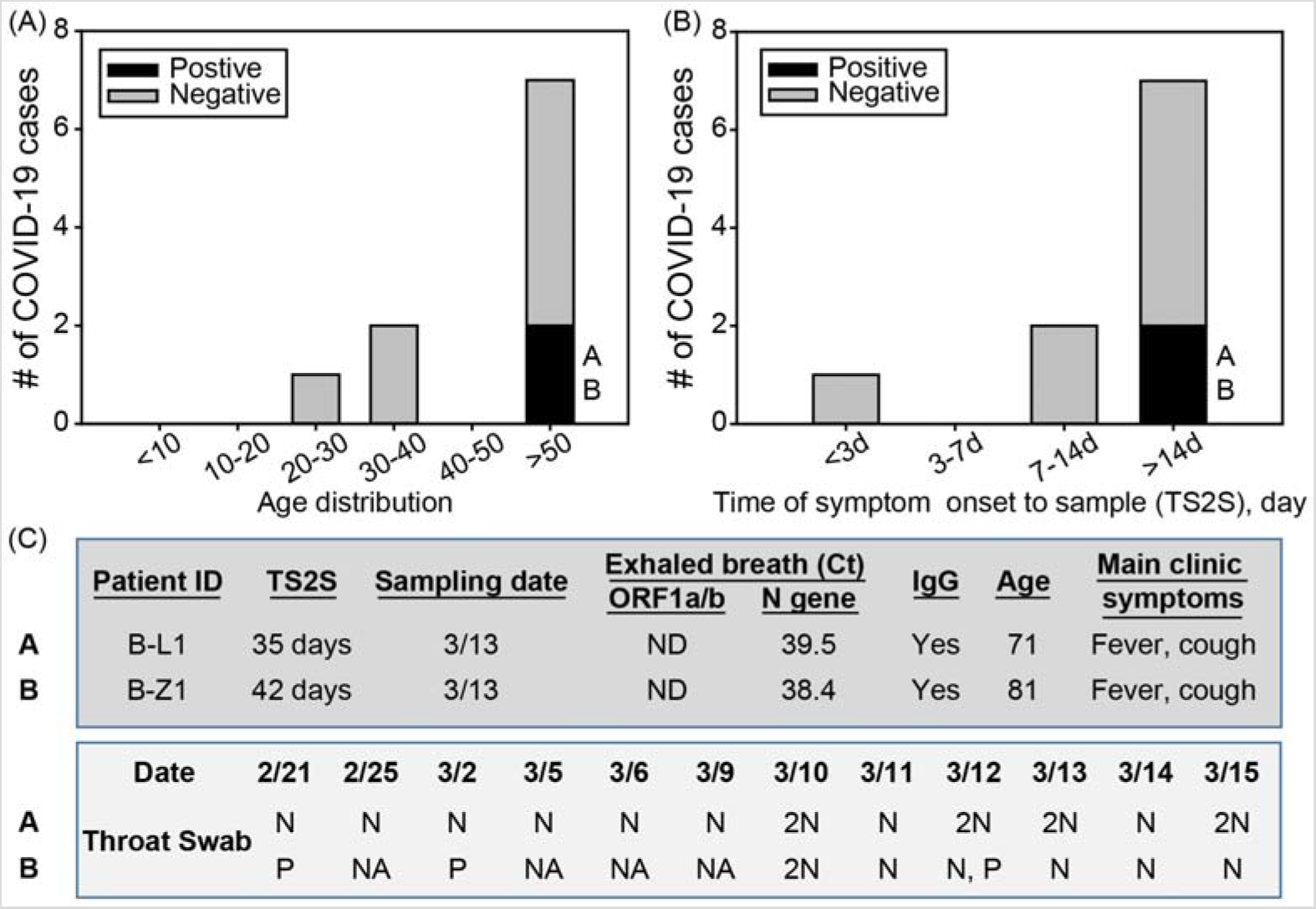
SARS-CoV-2 detection from EBC samples from the 10 recovering COVID-19 patients (Table S2); A) the recovering COVID-19 cases by age group; B) the recovering COVID-19 cases vs time of symptom onset to sample (TS2S) (day); C) medical records for the COVID-19 patients. 2N = two negative results on the same date. P = positive result; NA = no tests available; ND = not detected; N = negative result.

From the 318 surface swabs collected, four samples tested positive for SARS-CoV-2(Fig. 2). Two positive surface swab samples (Fig. 2B) were detected from the medical-staff-touching surface group. One was from a medical trolley D-SS-V8; the other (B-SS-D27), taken on 13 March, was from a medical instrument that had been used for monitoring the health condition of COVID-19 patient B-L2 who tested positive for SARS-CoV-2 on 9, 12, and 14 March. (Table S2). Another two positives included one (D-SS-V10) from the surfaces of the medical supply delivery window (Fig. S1D) installed between the general ward and the nursing center; another (D-SS-T8) from the shoe cabinets. Surprisingly, no positive samples were detected from the 158 patient-frequently-touched surfaces such as cell phones, door handles, patients’ hands, or even surfaces of the masks worn by the patients (D-Y1, D-Y2) (Table S4). In line with the surface observation, the EBC samples of the patients who were wearing the tested masks also tested negative (Table S3). None of the 57 hospital floor samples, 21 surface samples from clean areas, and 16 samples from other hospital surfaces tested positive for the virus. Unexpectedly from the common belief, the observed overall positive rate for the surface swab samples was strikingly low, only 1.3% (N=318).

**Fig. 2.**
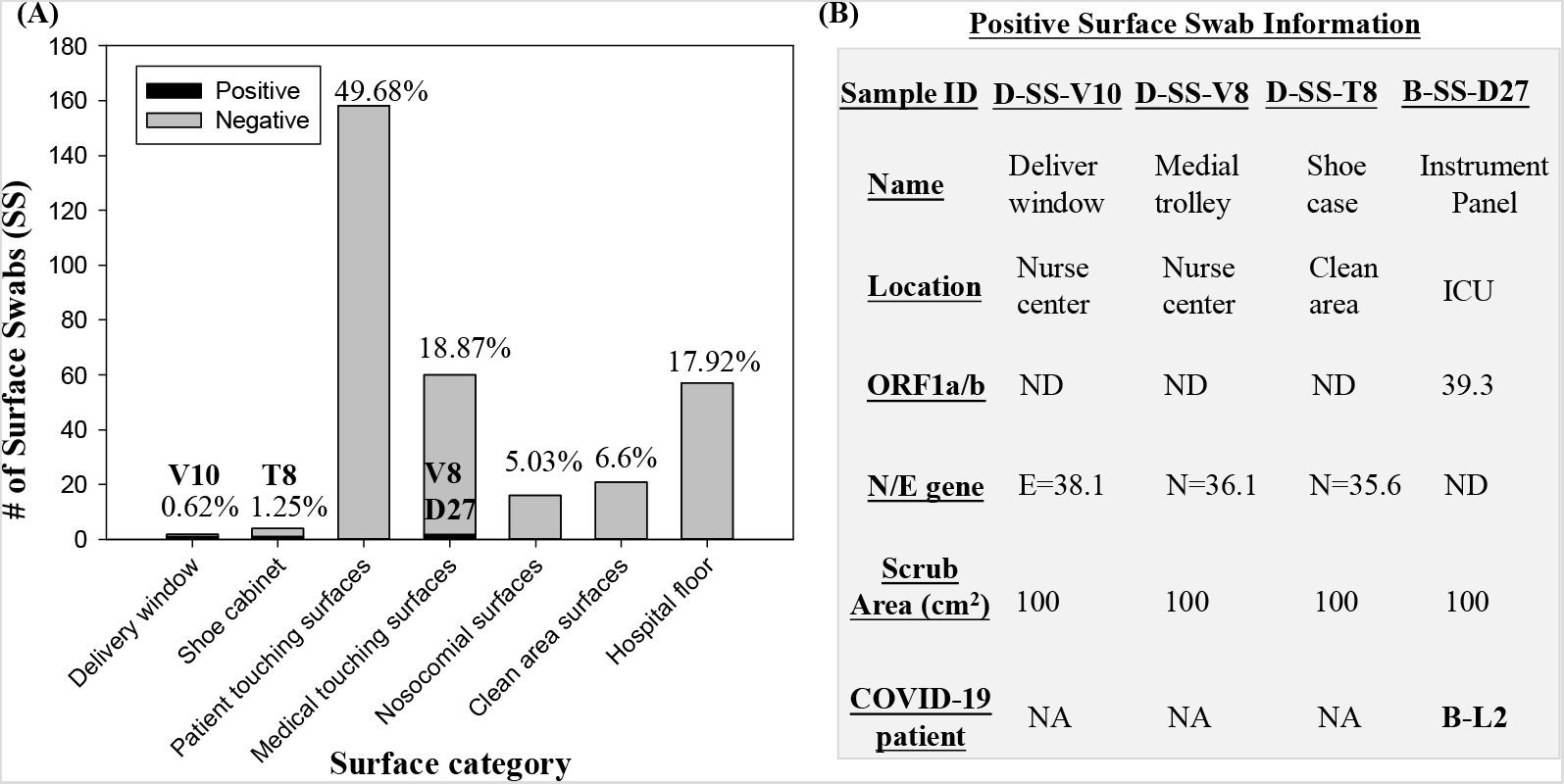
Detection of SARS-CoV-2 from surface swabs collected from seven categories of various sample collection locations: A) statistics for both positive and negative surface samples (total numbers and percentages); B) associated information of the detected positive surface samples. ND = not detected; NA = no tests available.

**Fig. 3.**
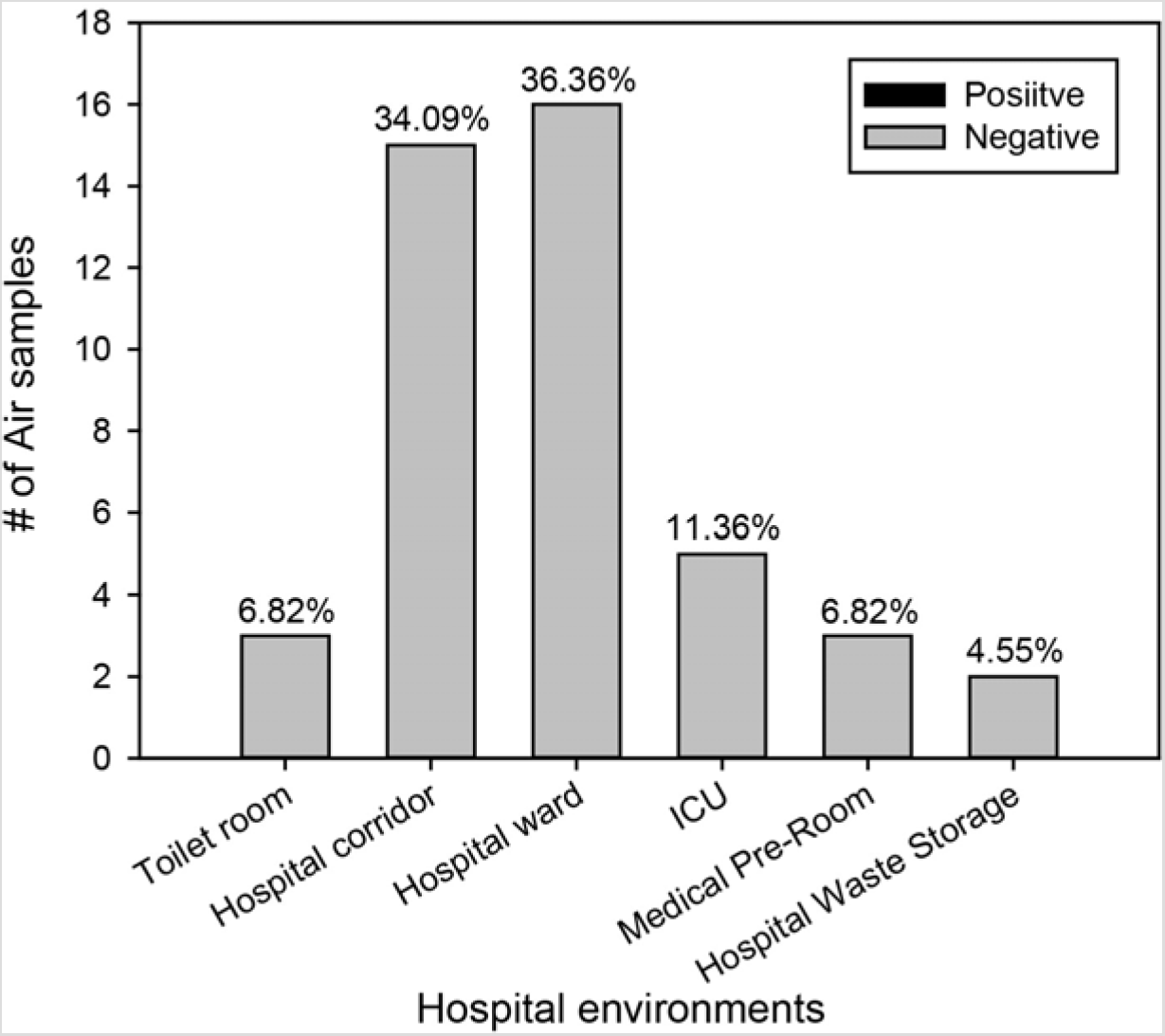
Detection of SARS-CoV-2 in air samples collected from six categories of different sampling locations. Percentages in the figure correspond to the fraction of each category of the total number of air samples.

None of the 44 air samples, including those collected using the robot (Video S1), tested positive for SARS-CoV-2 using RT-PCR. Even in the five intensive care unit (ICU) air samples, no SARS-CoV-2 was detected. Using the described chip technology, six air samples (A-Air-1–6, A-Air-Un1) (Table S5) were re-tested for both target SARS-CoV-2 genes; all of them were confirmed negative.

The highest positive rate for SARS-CoV-2 was observed from the exhaled breath samples (20%), followed by the surface swabs (1.3%) and air samples (0%). Surprisingly, two recovering COVID-19 patients (B-L1 and B-Z1), who repeatedly tested negative with their throat swabs, were observed to emit SARS-CoV-2 via breathing. The emission rate was estimated to be around 1400 copies/ min via breathing into the air. Another work reported that two COVID-19 cases (one without clinical symptoms), who repeatedly tested negative using throat swabs, tested positive for SARS-CoV-2 with their bronchoalveolar lavage fluid samples^12^. These findings further support our observation that SARS-CoV-2 could be emitted from the lungs via breathing, presenting a significant health risk for surrounding people and environments. This highlights a serious hidden airborne infection risk from the recovering COVID-19 patients, who were ready for hospital discharge using throat swab sample, but failed the EBC test. However, throat swabs and EBC specimens might work differently with different COVID-19 patients in terms of SARS-CoV-2 screening. COVID-19 diagnosis efficacy could be further improved by using EBC specimens or additional techniques such as CT scans as a complement to current throat swab testing by RT-PCR. Because of resource constraints, we did not study the viability and biological integrity of breath-borne SARS-CoV-2. Nonetheless, one study showed that aerosolized SARS-CoV-2 could remain viable in the air for up to 3 hours^13^. Logically, breath-emitted SARS-CoV-2, in a more favourable form of aerosolization, would be active in the air for a similar time scale, thus capable of facilitating the spread of the COVID-19. Information about airborne emission and the viability of SARS-CoV-2 emitted by patients is critical to understanding COVID-19 transmission. As the first effort reported, the present work revealed the SARS-CoV-2 presence in exhaled breath from recovering COVID-19 patients to be discharged, highlighting the underlying risk (20% failure rate) of the existing protocol in discharging COVID-19 patients.

Contrary to the current belief that direct surface contact represents a major route for COVID-19 transmission, we detected a very low positive rate (1.3%) for surface swabs (N=318) from various settings in the four Wuhan hospitals. This finding implies that direct surface contact, even in high risk areas in the environments studied, may not represent a major route of COVID-19 transmission. Using conventional RT-PCR, SARS-CoV-2 levels were shown below the detection limits for all 44 air samples collected (Video S1) in various hospital environments. The observed low positive rates both for the air and surfaces were a collective consequence of a number of factors. Firstly, virus emission dynamics from COVID-19 patients – when, how, where, and at what rate patients emit SARS-CoV-2 – are still largely unknown. The observed virus emission by at least one patient was not continuous (patient B-Z1, Fig 1); it may strongly dependent on the patient’s activities, e.g., coughing, sneezing, talking, or lung self-cleaning during the day. However, such activities, which are difficult to document, may have occurred before sample collection. Secondly, the hospitals applied disinfectants three times a day, possibly inactivating the virus and its RNA segments. Furthermore, all COVID-19 patients were required to wear a mask while in the hospital, reducing the release of the virus into the air or onto surfaces in the hospital environment. Lastly, natural air ventilation via open-windows (every room has at least one window with an outside wind speed of up to 1.6–3.3 m/s) allowed airborne virus to be rapidly diluted away. Since the time this study was conducted, no infections of medical staff have been reported from these four hospitals. The results from air samples indicate that the bio-safety control measures adopted by the hospitals for the air space were fairly effective. Nonetheless, our surface swab data suggest that certain surfaces frequently touched by the medical staff should be regularly disinfected to further lower related infection risks. The reported virus levels could have been underestimated by the RT-PCR method, which was previously reported to have a detection limit of 100 SARS-CoV-2 RNA copies/µL^14^. While low infection risks were shown for surfaces and air in the evaluated hospitals, our data reveal a critical need to revisit current hospital discharge guidelines to minimize the public risk.

## Methods

### Details of COVID-19 patients and the four hospital configurations

We recruited a total of 13 patients from four hospitals (Table S1, S2) in Wuhan, China for this study; and ten of whom were recovering COVID-19 patients ready for discharge based on repeated throat swabs tests (Table S2). The hospital wards had windows which were open during the sample collection. Male patients accounted for 60% of the 10 COVID-19 patients. Statistical distributions of age, severity of disease (the highest recorded during the disease course), and the time from the onset of symptoms to sample collection are shown in Fig. S1A, B (Table S2). The COVID-19 patients were aged from 29 to 81, with 70% of the patients older than 50, as shown in Fig. S1A; half had experienced severe symptoms during the disease course (Fig. S1B). Medical records for all patients were collected at the time of the sample collection (Table S2). The recruitment was conducted from 26 February –25 March, 2020; and the ethics involving non-invasive exhaled breath collection from patients was approved and waived due to the emergency situation by the Ethics committee of Jiangsu Provincial Center for Disease Prevention and Control. Fig. S1 C, D, E and Fig. S2, and S3 show representative hospital layouts, including designated areas, windows, natural air flow directions, and walking directions from the entry to the exit. The hospitals (Table S1) were clearly divided into different functional areas, including nursing center, clinical observation room, doctors’ office, restroom, work shift area (disinfection area for medical staff before exiting), and clean areas as indicated in Fig. S1. The usages of these areas were restricted to their designated hospital functions. The meteorological parameters and other information about ambient pollutants during the monitoring campaign are listed in Table S6. The average wind was observed to be category II, which corresponds to a wind speed of 1.6–3.3 m/s. The hospital rooms in four hospitals of Wuhan used a natural ventilation.

### Exhaled breath sample collection

Exhaled breath condensate (EBC) samples were collected from 13 recruited patients (10 recovering COVID-19 patients, and three with influenza symptoms who tested negative for SARS-CoV-2), their using a BioScreen II device (Beijing dBlueTech Inc., Beijing, China) following the protocols provided by the manufacturer (Fig. S4). To avoid saliva contamination, a long straw was used to allow the patient to breathe into a tube that was electrically cooled. The EBC sample of one non-COVID-19 patient (C-Y1) was collected twice to confirm the result. EBC sample volumes of approximately 300–500 μL were obtained for all patients. The samples were immediately pipetted into a corning tube and transported to the laboratory for SARS-CoV-2 analysis. The clinical information for non-COVID-19 patients is listed in Table S2. The throat swabs of the recovering COVID-19 patients were all tested negative for SARS-CoV-2 by the hospital (Table S2) before their EBC samples were collected. Examples of the EBC sample collection points in four different hospitals are shown in Fig. S2 and S3. A total of 14 EBC samples were collected (see Table S3 for detailed information).

### Surface swab sample collection

To evaluate environmental contamination with SARS-CoV-2, swab samples were collected from surfaces associated with the COVID-19 patients and medical staff, and from many other surfaces inside the four hospitals in Wuhan. Specifically, a wet cotton swab was used to scrub a surface (an area of 10 cm × 10 cm or 5 cm × 5 cm) of objects in the hospital environment and from personal items of the patients. The surface swab samples were deposited in the virus collection liquid (Jiangsu Kangjian Medical Supply, Inc, Nanjing, China), and then transported to the laboratory and stored at –20°C for SARS-CoV-2 analysis. A total of 318 surface swabs were collected. Details are listed in Table S4.

### Air sample collection

To examine environmental bio-safety, air samples were collected from the corridors, hospital waste storage rooms, ICU rooms, toilets, medical preparation rooms, clinical observation rooms, and general wards of four hospitals in Wuhan, China. The air samples were collected using impinger samplers (WA-15, WA-400; Beijing dBlueTech, Inc., Beijing, China) (examples for onsite sampling are shown in Fig.S5). The WA-15 sampled at a flow rate of 15 L/min, while the WA-400 sampled at 400 L/min. For corridor spaces or naturally ventilated environments, the WA-400 was installed on a robot for air sampling (Fig. S5B), while for semi-enclosed environments such as toilets or ICU rooms, the WA-15 was used for sampling (Fig. S5A). The robot was programmed to move along pre-determined routes inside the hospital (Fig. S5C; Video S1). In each case, air was sampled into 3 mL of virus sampling liquid (Jiangsu Kangjian Medical Supply, Inc., Nanjing, China) for 40 min; after the sampling, about 1.5–2 mL collection liquid remained due to the evaporation. The collected air samples were transported to the laboratory for SARS-CoV-2 analysis. The air samples collected using the robot comprised air from different areas of the interior hospital corridor, and thus were more representative than those from a stationary sampling. Representative air sample collection points are also shown in Fig. S2, and S3. A total of 44 air samples were collected; detailed information is listed in Table S5.

### SARS-CoV-2 analysis for the air samples, exhaled breath condensate samples and surface swabs using RT-PCR and loop-mediated isothermal amplification (LAMP)

The collected samples (200 μL taken from the collection liquid) went through several steps for SARS-Cov-2 detection. First, an automated nucleic acid extraction device (NP968-S, Xi’an Tianlong Sci &Tech Co., Ltd, Xi’an, China) and an RNA extraction kit (Jiangsu Bioperfectus Technologies, Nanjing, China) were used to extract SARS-CoV-2 RNA, achieving a final sample RNA suspension of 70–80 μL. SARS-CoV-2 detection was then performed using RT-PCR (BioRAD CFX96 Real-Time System C1000 Thermal Cycler, Hercules, California) together with a detection kit (Jiangsu Bioperfectus Technologies) except the sample D-SS-V10 (N, E and RdRP genes analyzed) under the following cycle conditions: 50°C for 10min, and 97°C for 1min, followed by 45 cycles of 97°C for 5s, and 58°C for 30s. The reaction mixture included 7.5 μL of nucleic acid amplification mix, 5μL of Taq EnzymeMix, 4 μL of SARS-CoV-2 reaction mix, 3.5 μL of RNA-free H_2_O, and 5 μL of sample RNA. In accordance with the instructions, for cycle threshold (Ct) values of less than 37, and those less than 40 but greater than 37, with an “*S”* shape amplification curve, the corresponding samples were treated as a positive. The RT-PCR detection limits using various primer sets were shown to be around 100 SARS-CoV-2 RNA copies/μL; those obtained by the Chinese CDC were lower (below 50 RNA copies/μL) for the N gene with a maximum Ct value of approximately 39.5^14^. The primer set was not disclosed for the kit used here by the company. Some of the collected samples were retested for SARS-CoV-2 for both N and ORF1a/b genes using the LAMP chip technology (Beijing CapitalBio Technology Co., Ltd., Beijing, China) following the manufacturer’s instructions. In addition to SARS-CoV-2, this technology can also detect 18 other viruses. All samplings were performed using single-use consumables and deionized (DI) water served as the negative controls.

## Data Availability

All data are included in the manuscript and supporting information files.

## Acknowledgements

This research was supported by a National Natural Science Foundation of China (NSFC)grant (22040101) (PI: M. Yao), the Chinese Academy of Engineering Grant(2020-ZD-15) dedicated to the COVID-19 pandemic. This work was also partially supported by the NSFC Distinguished Young Scholars Fund Awarded to M. Yao (21725701), and Scientific Research Fund of Jiangsu Provincial Health Committee(S2017002).

## Author Contributions

M.Y., Y.Z., W.L., and B.Z. conceptualized the study design, provided resources, and managed the project; L.Z.,X.Z.,Y.L.,M.D.,B.S.,Y.J.,K.Z.,J.H., and N.Y. collected samples and performed the laboratory tests; M.Y. provided the sampling and analysis devices; M.Y., L.Z.,Y.Z., W.L., B.Z., L.M., S.A.G., P.B., and R.C.F. analysed the data; M.Y., L.Z.,Y.Z.,W.L.,L.M.,S.A.G., P.B., and R.C.F. interpreted the results; M.Y. wrote the initial drafts of the manuscript, and X.L., L.Z., and H.C. assisted the manuscript figure preparation and references; M.Y., L.Z.,Y.Z.,W.L., B.Z., M.H., L.M., S.A.G., P.B., R.C.F., X.L., L.Z., H.C., Z.D., and Y.X. revised and commented the manuscript. All authors read and approved the final manuscript.

## Declaration of interests

We declare no competing interests.

## Additional Information

Supplementary Information is available for this paper.

## Notes

### Competing Interest Statement

The authors have declared no competing interest.

### Funding Statement

This research was supported by the National Natural Science Foundation of China grant (22040101), the Chinese Academy of Engineering Grant (2020-ZD-15 ) dedicated to the COVID-19 pandemic. This work was also partially supported by the NSFC Distinguished Young Scholars Fund Awarded to M. Yao (21725701), and Scientific Research Fund of Jiangsu Provincial Health Committee(S2017002).

### Author Declarations

The recruitment was conducted during the period of 26 Feb -25 March, 2020; and the ethics involving patients' exhaled breath non-invasive collection was approved and waived due to the emergence matter by the ethics committee of Jiangsu Provincial Center for Disease Prevention and Control.

